# Characterizing the Short- and Long-Term Temporal Dynamics of Antibody Responses to Influenza Vaccination

**DOI:** 10.1101/2025.02.26.25322965

**Authors:** Aaron Lane, Huy Q. Quach, Inna G. Ovsyannikova, Richard B. Kennedy, Ted M. Ross, Tal Einav

## Abstract

Most influenza vaccine studies evaluate acute antibody responses 1 month post-vaccination, leaving long-term immunity poorly understood. Here, we performed a combined analysis of 14 large-scale vaccine studies and conducted two new studies mapping antibody responses in high resolution from their inception out to 1 year post-vaccination. Vaccine antibody responses were classified as *weak* (<4x fold-change at 1 month and 1 year), *transient* (≥4x at 1 month, <4x at 1 year), or *durable* (≥4x at 1 month and 1 year). Surprisingly, >50% of vaccine recipients were weak across seasons, age groups, sexes, pre-vaccination titers, and high or standard vaccine doses. Peak fold-change at 1 month post-vaccination was strongly associated with the long-term response, with most transient responders achieving a maximum fold-change of 4x, while most durable responders reached ≥16x, with both groups maintaining these titers for 2 months (10-75 days post-vaccination). Using the weak, transient, and durable trajectories, a single time point early in the response (days 7-8 or 21) predicted an individual’s response out to 1 year post-vaccination. These results demonstrate that influenza vaccine responses range from little-to-no response to eliciting strong-and-durable immunity, highlighting the stark heterogeneity that is consistently seen across influenza seasons.

## Introduction

While influenza vaccination provides population level immunity, individual responses vary widely and this heterogeneity remains poorly understood. The same vaccine may elicit a strong response in some, but little-to-no response in others, and we currently lack tools to predict who will respond weakly or estimate how alternate formulations (*e.g.*, high dose, adjuvanted, or recombinant vaccines) would improve such responses. As a result, influenza vaccine effectiveness remains around 20-50%, even in the majority of seasons when the vaccine strain matches the circulating strain.^1,2^

While many studies have examined the magnitude of the vaccine response, relatively few have quantified how vaccine durability varies across individuals. Most data on the long-term influenza response comes from test-negative case-control studies of influenza vaccine effectiveness^3^ ‒ where individuals are PCR tested at a single time point ‒ which showed that effectiveness can drop to 0% within 3 months post-vaccination.^4,5^ Yet these analyses only estimate the average vaccine effectiveness across the population, and they rarely track individuals over time nor capture population-level heterogeneity. For example, the above statistics could reflect a scenario where most of the population shows little response to vaccination, while the rest mount a strong, long-lasting response. Indeed, given that in most individuals influenza-specific B cells are not recruited to the bone marrow as long-lived plasma cells following vaccination,^6^ the few durable responses would be swamped out when examined at the population-scale.

To address this shortcoming, some influenza vaccine studies track individual antibody responses longitudinally, relying on the known association between strong antibody responses and protection.^7,8^ However, even with person-level data, many only report the average response across subjects. Additionally, most studies only measure antibody levels pre-vaccination and 1 month post-vaccination. The few studies extending beyond this time frame corroborated the vaccine effectiveness results above, reporting that the antibody response peaks at 1 month post-vaccination before returning to baseline levels within 3-6 months post-vaccination,^9,10^ suggesting that the influenza vaccine does not induce long-term durability.

Here, we collected data from influenza H3N2 vaccine studies from multiple seasons and conducted two new vaccine studies using samples from 2018 and 2022 to examine individual-level responses. Despite changes in the vaccine strain across seasons, each person’s antibody response consistently fell into a few phenotypes, allowing us to combine data from these large-scale studies. In doing so, we determined the dynamics of the antibody response with high resolution, quantifying its onset, peak size, duration, and long-term decay out to 1 year post-vaccination.

Prior work noted that the influenza vaccine can elicit a transient antibody response in some and a durable response in others (also called temporary and persistent responses^11^), yet to our knowledge the frequency of these phenotypes has not been quantified. In addition to transient and durable responses, we also define a weak phenotype where individuals exhibit little-to-no antibody response post-vaccination, and surprisingly, we find that this phenotype is by far the most common across all seasons. The predominance of these weak responders suggests that the average statistics computed in the past, while correct for the population as a whole, have greatly underestimated the effectiveness of the influenza vaccine in some individuals and greatly overestimated it in others.

## Results

To determine the short- and long-term dynamics of the influenza H3N2 vaccine response, we performed a literature search and found 9 large-scale influenza studies for our initial analysis, each assessing 41-320 individuals across 3-6 time points using hemagglutination inhibition titer (referred to as HAI hereafter) (**Table 1**, white background). The combined dataset contained ∼1300 antibody responses covering ∼200 different days of the post-vaccination response and six different H3N2 vaccine strains, and to our knowledge these studies constitute the most comprehensive coverage of both short- and long-term influenza vaccine response dynamics. For each study, we analyzed the fold-change in post-vaccination (post-vac) HAI relative to day 0 for that season’s H3N2 vaccine strain (absolute post-vac HAI shown in **Fig S1**).

**Table 1.** Large-scale influenza vaccine studies analyzed. Each measured time point is separated by a comma, while exact time ranges (when reported) are indicated by a hyphen. (# Data Points) = (# Sera)×(# H3N2 Variants)×(# Times Measured)-(# of Missing Measurements). White backgrounds denote prior studies while gray backgrounds represent new studies introduced in this work. Three additional studies without variants that were included in this work are listed in **Table S2**.

| Year + Name of Study* | Number of Data Points | Number of Sera | Number of H3N2 Variants | Times Measured | H3N2 Vaccine Strain (Formulation) |
| --- | --- | --- | --- | --- | --- |
| 2014 Hinojosa<sup>12</sup> | 1968 | 41 | 16 | [0, 28, 365] | A/Texas/50/2012 (Fluzone or FluMist) |
| 2016 Fox<sub>Nam</sub> (Ha Nam)<sup>9</sup> | 23862 | 97 | 41 | [0, 4, 7, 14, 21, 280] | A/Hong Kong/4801/2014 (Vaxigrip) |
| 2016 Fox<sub>HWC</sub> (Health Care Workers)<sup>9</sup> | 5292 | 49 | 36 | [0, 21, 224] | A/Hong Kong/4801/2014 (Fluarix) |
| 2016 UGA<sup>13</sup> | 5202 | 102 | 17 | [0, 21, 186-389] | A/Hong Kong/4801/2014 (Fluzone) |
| 2017 UGA<sup>13</sup> | 8100 | 150 | 18 | [0, 21, 207-430] | A/Hong Kong/4801/2014 (Fluzone) |
| 2018 UGA<sup>13</sup> | 4860 | 162 | 10 | [0, 28, 263-490] | A/Singapore/INFIMH-160019/2016 (Fluzone) |
| 2019 UGA<sup>13</sup> | 7680 | 320 | 8 | [0, 28, 238-429] | A/Kansas/14/2017 (Fluzone) |
| 2020 UGA<sup>13</sup> | 5250 | 250 | 7 | [0, 21-71, 311-444] | A/Hong Kong/2671/2019 (Fluzone) |
| 2021 UGA<sup>13</sup> | 3255 | 155 | 7 | [0, 21-43, 323-447] | A/Tasmania/503/2020 (Fluzone) |
| 2022 UGA | 4571 | 168 | 7 | [0, 19-75, 76-108, 284-399] | A/Darwin/9/2021 (Fluzone) |
| 2018 Kennedy | 632 | 212 | 1 | [0, 8, 28] | A/Singapore/INFIMH-160019/2016 (Fluzone or Fluad) |

\* Each year represents the date when a vaccine study was conducted, not when the corresponding manuscript was published.

### The power of repeat vaccinations: Assessing long-term timepoints 1 year post-vaccination

Among the studies in **Table 1**, only two (2016 Fox_Nam_, 2016 Fox_HCW_) explicitly measured long-term responses more than 180 days post-vac. However, such long-term responses can be determined for individuals who participated in consecutive studies, where their pre-vaccination (pre-vac) titers in the following season quantify their 1 year post-vac response in the current season (**Fig S2**). Crucially, when the vaccine strain changed, as happened every 1-2 years for H3N2, this association was only possible in studies that measured the HAI of prior vaccine strains (**Fig S2**). By focusing on such studies, we determined a long-term time point for 1180 sera between days 180-500 post-vac, providing greater insight into vaccine durability at both individual and population levels.

### Most influenza vaccine responses at 1 month post-vaccination are weak, but strong responders are equally likely to be transient or durable

Given the heterogeneity of responses across individuals and across seasons, we first examined the magnitude of the peak response 1 month post-vac, which is the only post-vac time point measured in all studies. Surprisingly, the geometric mean (geomean) fold-change of all 1326 individuals was 2.9x at 1 month (95% CI: 1.9-4.7x across seasons, **Table S1**), which in most seasons fell below the 4x fold-change that often represents an appreciable “seroconverted” vaccine response.

However, this average 2.9x increase ignores substantial variability across individuals, particularly at the peak (1 month post-vac) and long-term (1 year post-vac) time points. At 1 month post-vac, 59% of individuals in all studies exhibited little-to-no response (≤2x, the noise limit of the HAI assay). In the opposite limit, 14% of all subjects had a large fold-change ≥16x. At the long-term time point 1 year post-vac, 79% of all individuals returned to ≤2x of their baseline, while a surprisingly large 21% maintained ≥4x fold-change.

To better capture this variability, we classified individuals based on their peak and long-term HAI. Responses were classified as *strong* if HAI fold-change ≥4x at 1 month against the vaccine strain and at least one additional variant (to avoid misclassification due to experimental noise, since the HAI dynamics of variants closely match the vaccine strain’s dynamics [**Fig S3**]); all other responses were classified as *weak* (**Fig 1A**). Strong responses were subcategorized as *durable* if they maintained ≥4x fold-change against the vaccine strain and one other variant at their long-term time point (180-500 days post-vac), and otherwise categorized as *transient* if their long-term response fell to <4x (**Fig 1A,B**).

**Figure 1.**
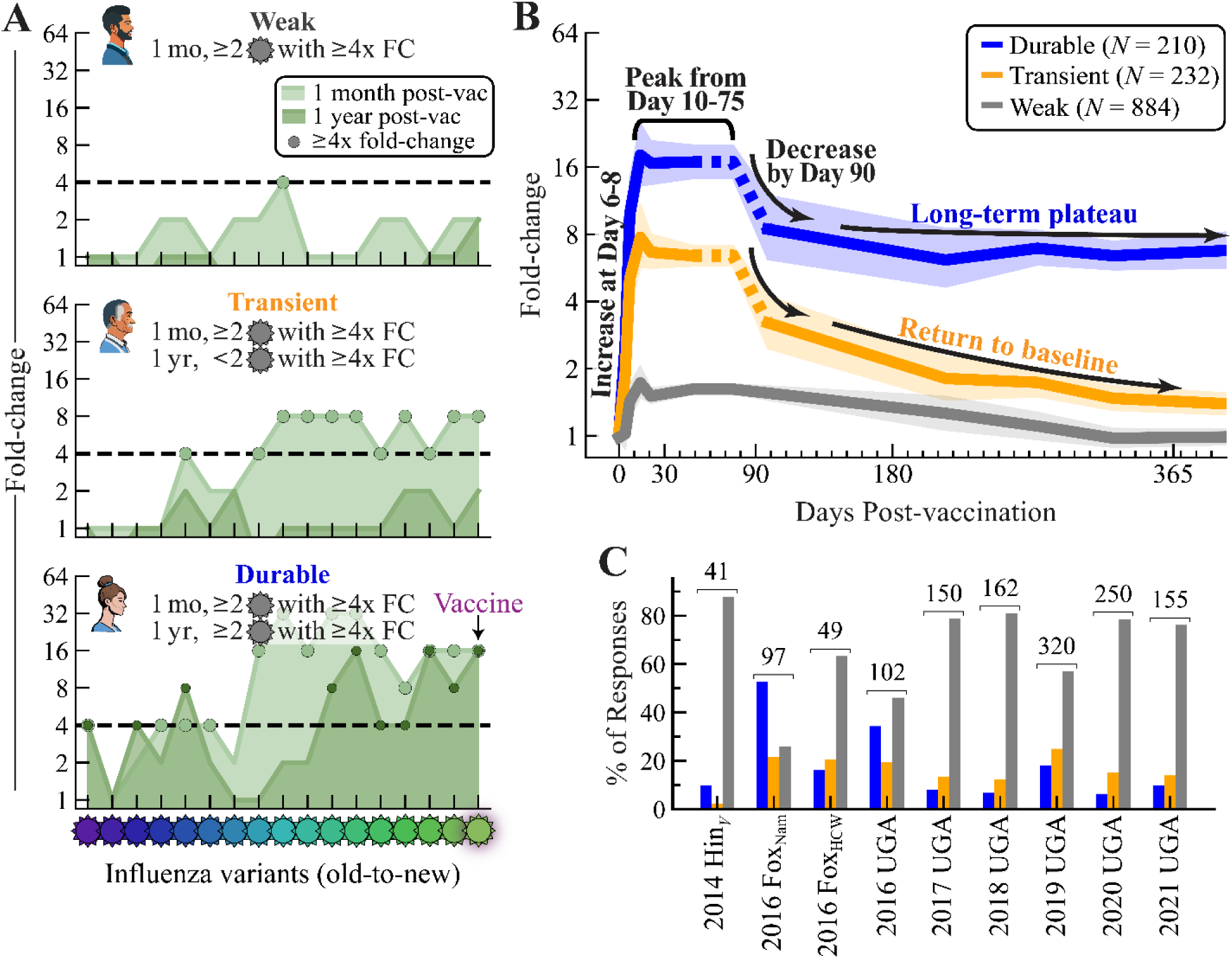
Durable, transient, and weak phenotypes of post-vaccination HAI fold-change. (A) Three representative subjects from the 2016 UGA study, grouped according to their fold-change=(HAI titer post-vaccination)/(HAI titer at day 0) as durable (≥4x fold-change for ≥2 variants at both 1 month and 1 year), transient (≥4x fold-change for ≥2 variants at 1 month but *not* at 1 year), and weak (all remaining responses). (B) Geometric mean of *N*=1326 responses from the 2014-2021 influenza seasons at all post-vac time points (**Methods**), grouped as durable (blue), transient (gold), and weak (gray) phenotypes. Lines show the mean and 95% confidence interval. Dashed segments for durable and transient responses were inferred from single datasets (**Methods**). (C) Fraction of responses in each category across datasets in **Table 1** (white background). The total number of participants is shown above each dataset.

The weak, transient, and durable phenotypes were seen in every study, although transient and durable responders may be rare (**Fig 1C**). Across all studies, 67% (884/1326) of vaccine responses were weak (**Table 1**, white background). Of the strong responses, 53% (*N*=232) were identified as *transient* and the remainder (*N*=210) were *durable*. Despite season-to-season variability, most individual studies followed this same trend, with two notable exceptions being 2016 Fox_Nam_ and 2016 UGA that had the lowest fractions of 26% and 46% weak responders, respectively (**Fig 1C**). All other studies had ≥50% weak responders.

To determine which factors contribute to this large fraction of weak responders, we stratified subjects by sex, vaccine dose (standard vs high-dose, Fluzone only), age, and pre-vac HAI. Sex and vaccine dose had little effect, with each category retaining 65-70% weak responders (**Fig 2A,B**). Compared to older adults (30-64 years old, 54-61% weak), the elderly (65+ years old, 67% weak) were slightly more prone to the weak phenotype (**Fig 2C**). Surprisingly, young adults (20-29 years old) and children (5-19) had the highest likelihoods (73% and 77%, respectively) of having weak responses, countering the expectation that they elicit the strongest responses (**Fig 2C**).

**Figure 2.**
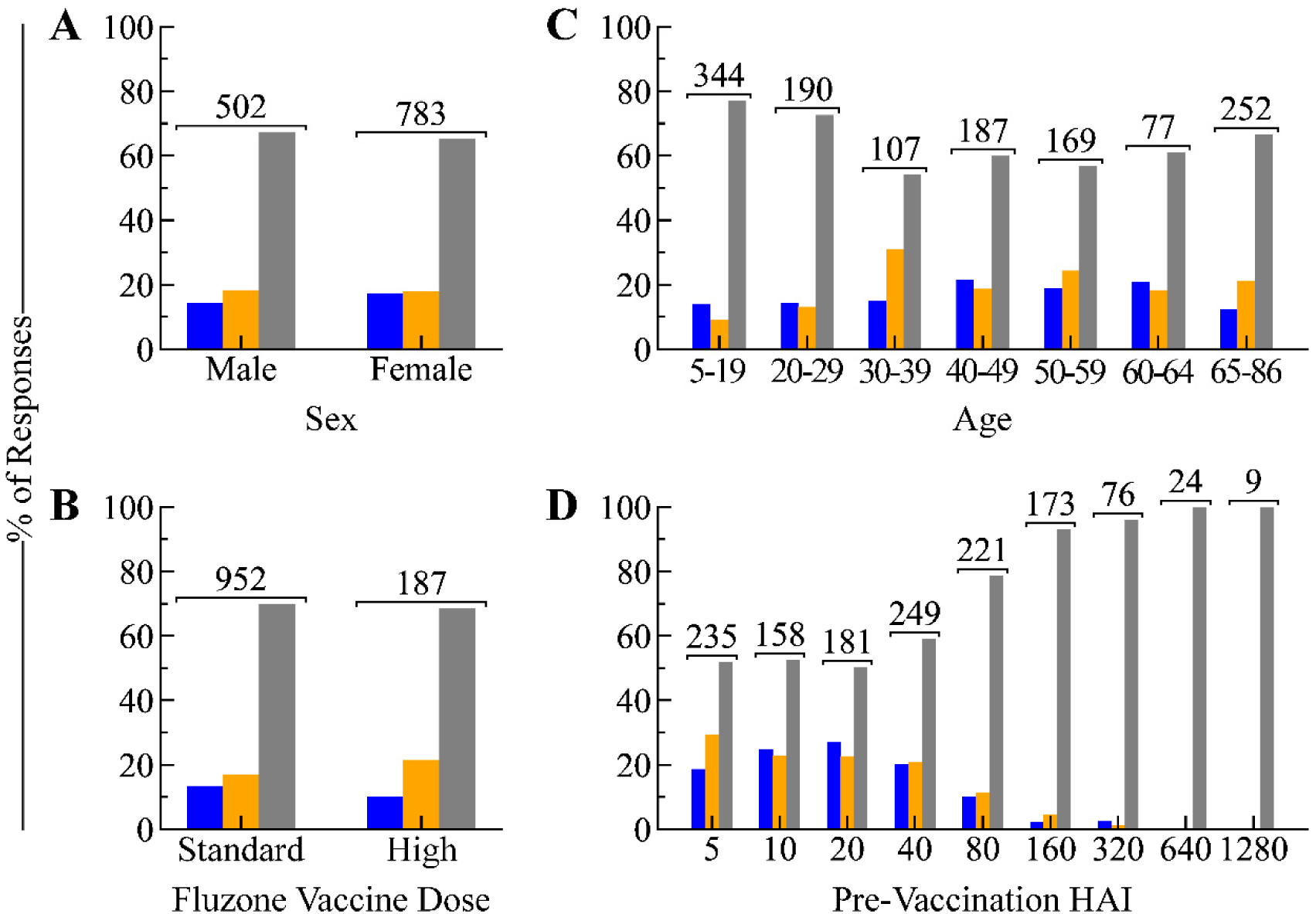
Fraction of weak, transient, and durable responses across individual factors. Fraction of responses in each phenotype across (A) sex, (B) vaccine dose in Fluzone studies [high-dose has 4x more antigens than the standard dose], (C) age, and (D) pre-vaccination titers. The total number of participants is shown above each group.

Pre-vac titers had a more pronounced effect on their phenotype compared to the above factors. Pre-vac HAI≤40 led to 50-60% weak responders, but the odds of being weak drastically increased for pre-vac titers of 80 (79%), 160 (93%), and 320 (96%), with the few individuals with higher pre-vac titers having a 100% chance of being weak (**Fig 2D**). Thus, a large pre-vac titer skews the response towards the weak phenotype, although we note that most individuals have HAI≤40 before vaccination.

The following three sections present a series of vignettes that examine the dynamics of the weak, transient, and durable phenotypes in temporal order, starting from the initiation of the antibody response (days 0-14) and ending with the long-term response (1 year).

### HAI begins to increase 7 days post-vaccination for both transient and durable responders

To determine when the antibody response starts to increase post-vaccination, we calculated when fold-change reached ≥4x in two studies that measured the early vaccine response (days 0-14). In the first dataset, 2016 Fox_Nam_, sera were collected at days 0, 4, 7, 14, 21, and 280. The latter two time points categorized responses as weak, transient, or durable, while the three early time points determined when the response began. 81% of transient and 82% of durable responders achieved ≥4x fold-change by day 7, with linear interpolation suggesting that 50% of transient and durable responders achieve 4x fold-change around day 6 (**Fig 3A,B**). At day 7, durable responders already exhibited a slightly larger geomean fold-change (9.2x, 95% CI: 6.3-13.6) than transient responders (5.0x, 95% CI: 3.6-7.2x), with both groups exhibiting far more fold-change than weak responders (1.4x, 95% CI: 1.2-1.7x) that never achieved ≥4x at any time point.

**Figure 3.**
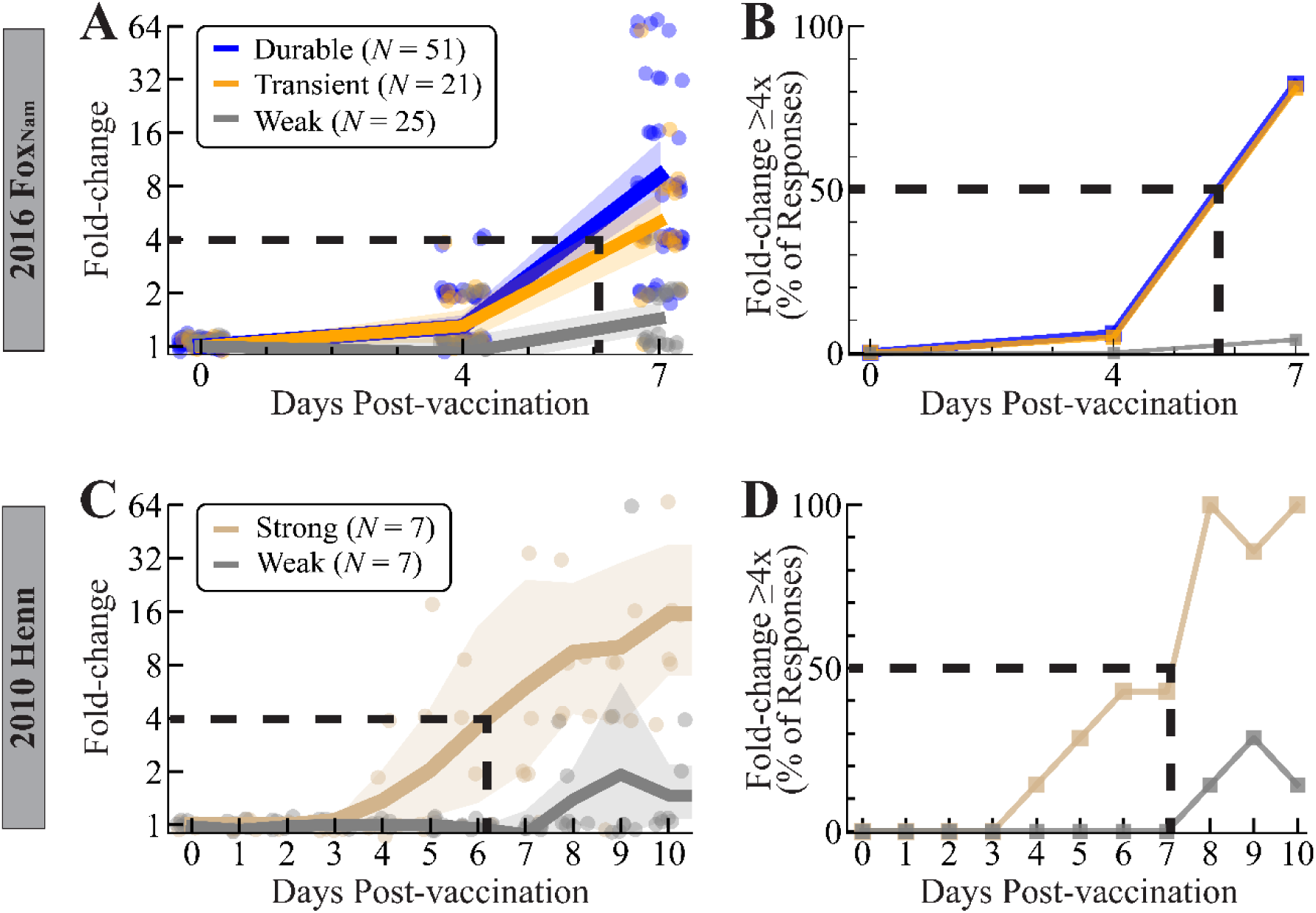
Time required for HAI titer to start increasing post-vaccination. (A,B) Responses from the 2016 Fox_Nam_ study^9^ showing (A) fold-change or (B) the percent of sera achieving fold-change ≥4x at days 0, 4, and 7 after vaccination with H3N2 A/Hong Kong/4801/2014. (C,D) Responses from the 2010 Henn study showing (C) fold-change or (D) the percent of serum responses achieving fold-change ≥4x after vaccination with H3N2 A/Perth/16/2009^14^ at days 0, 1, 2… 10. In Panels A and C, points show individual responses and lines show the geometric mean and 95% confidence intervals.

To examine the vaccine response in greater temporal resolution, we conducted a literature search and found the 2010 Henn study, where sera were collected daily (days 0, 1, 2…10) together with the peak response (day 21) (**Table S2**).^14^ Since this study lacked a long-term time point and only measured HAI against the 2010 vaccine strain (H3N2 A/Perth/16/2009), we only classified participants as weak (*N*=7) or strong (*N*=7) based on whether they achieve ≥4x fold-change at day 21 (**Methods**). Geomean fold-change across all 7 strong individuals reached 4x at day 6 (**Fig 3C**), with 6/7 consistently maintaining fold-change≥4x by day 8 (**Fig 3D**). Most weak responses never achieved ≥4x fold-change at any time point, and hence they were ignored. Collectively, these results suggest that individuals that will exhibit a strong response to the vaccine achieve a 4x rise around days 6-7.

### Strong responses achieve their peak fold-change 10 days post-vaccination and begin to wane by 90 days

While most influenza vaccine studies measure the peak antibody response 1 month post-vac, it remains unclear how early and how long this peak is sustained. To that end, we first quantified the magnitude of the peak at 1 month and then assessed its duration.

Across the 2014-2021 seasons, the weak, transient, and durable responses showed progressively larger fold-change at 1 month with geomeans of 1.6x (95% CI: 1.5-1.7x), 6.4x (95% CI: 5.9-7.0x), and 16.3x (95% CI: 14.2-18.8x), respectively (**Fig 4A, Methods**). Absolute HAI followed a similar trend with geomeans of 77, 112, and 313, respectively (**Table S3C**).

**Figure 4.**
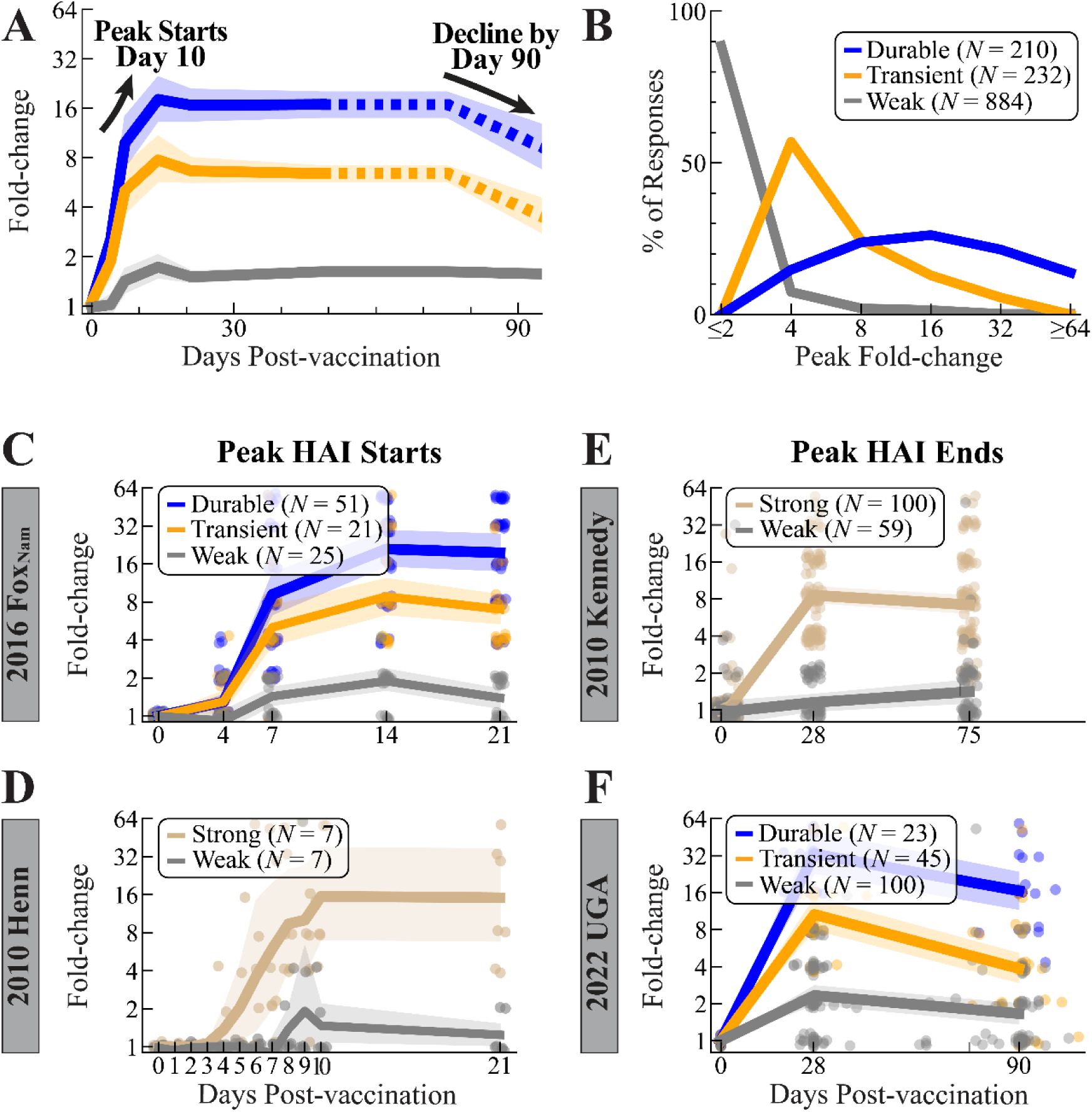
Timing and magnitude of the peak HAI titer response across weak, transient, and durable responders. (A) Fold-change for any participants measured between days 0-90 post-vaccination. Points show individual responses, lines show the cohort geometric mean fold-change and 95% confidence intervals. Dashed portions of these lines were derived from a single dataset ‒ the 2010 Kennedy study for day 75 and the 2022 UGA study for day 90.^15^ (B) Distribution of peak fold-change across each category. The start of the peak HAI titer response was determined using fold-change from (C) 2016 Fox_Nam_ and (D) 2010 Henn, while the end of the HAI titer peak was found using (E) 2010 Kennedy and (F) 2022 UGA (**Table 1**).

By definition, the fold-change of weak responders was tightly distributed at 1-2x (86%), except for the 14% that achieved ≥4x for the vaccine strain but not other variants. Though by definition transient responders (fold-change≥4x at 1 month post-vac) were bounded from below, they were also tightly distributed, with 81% ranging from 4-8x and only 19% exceeding 8x. In contrast, durable responses showed greater variability, with 81% exhibiting the most common 8-32x fold-change while 14% had ≥64x fold-change (**Fig 4B**).

Using two individual datasets with early time points, we scanned backwards in time from 1 month to determine when the peak response is achieved. In the 2016 Fox_Nam_ study, fold-change at days 21 and 14 were similar, with their combined geomean (FC_weak_=1.6x, FC_transient_=7.8x, FC_durable_=20.3x) matching the 1 month geometric mean from all studies. However, fold-change at day 7 was smaller (FC_weak_=1.4x, FC_transient_=5.0x, FC_durable_=9.2x) (*N*=97, **Fig 4C**; distributions across time points in **Fig S4**). Notably, the durable responders increased by >2x from day 7 (9.2x, 95% CI: 6.8-14.4x) to day 14 (21.0x, 95% CI: 13.6-26.8x). In contrast, the *N*=21 transient responders increased by less than the 2x noise limit (FC_day7_=5.0x, FC_day14_=8.8x), and even dropped modestly at day 21 (7.0x), making the onset of the HAI peak less clear for transient responders. Nevertheless, both these transient and durable responses suggested that peak HAI begins between day 7-14.

The *N*=7 strong responders from the 2010 Henn dataset (HAI at days 0, 1…10, 21) corroborated and refined this range. In this small dataset, fold-change at day 21 (FC_weak_=1.2x, FC_strong_=14.5x) matched that at day 10 (FC_weak_=1.3x, FC_strong_=17.7x) but was smaller at days 8 and 9 (FC_weak_=1.7x, FC_strong_=9.3x) (**Fig 4D**). In combination with the larger 2016 Fox_Nam_ study, peak HAI is reached 10-14 days post-vaccination.

We then turned to the other limit of the peak response and assessed when antibody responses start to decline. Since we found no vaccine studies that measured responses between 1 month and 1 year post-vac against multiple variants, we: 1) used an existing study that only measured HAI against the vaccine strain and 2) conducted our own vaccine study that also measured variants. The first study, 2010 Kennedy, measured responses at days 0, 28, and 75 post-vaccination against the vaccine strain alone (*N*=159, **Fig 4E**).^15^ As before, since this dataset lacked a long-term time point or additional H3N2 variants, we classified participants solely as weak (*N*=59) or strong (*N*=100) based on their day 28 response (**Methods**). Notably, fold-change at day 75 (7.2x, 95% CI: 6.0-8.5x) was comparable to fold-change at day 28 (8.6x, 95% CI: 7.4-10.1x) among strong responders, with 61% seeing no change in HAI (**Fig S4G**), suggesting that the peak lasts at least until day 75.

We also conducted a new vaccine study, 2022 UGA, that measured sera at days 0, 28, 90, and 365 (*N*=168, **Fig 4F**). To our knowledge, this is the first study that measures the short-term waning at day 90 and long-term response at 1 year against multiple variants. This study followed the same format as the prior UGA studies with the addition of this day 90 time point, administering that season’s Fluzone Quadrivalent vaccine (containing H3N2 A/Darwin/9/2022) (**Table 1**, gray background). As in all prior studies, most responders were weak (*N*=100), with fewer transient (*N*=45) or durable (*N*=23) responders. Compared to 1 month (geometric mean FC_weak_=2.3x, FC_transient_=10.7x, FC_durable_=33.0x), the transient and durable responders dropped by ∼2-fold at day 90 (FC_weak_=1.6x, FC_transient_=3.9x, FC_durable_=16.6x). Together with the 2010 Kennedy study, this suggests that peak HAI ends between days 76-90.

As in all vaccine studies, a potential confounder is that individuals may experience breakthrough infections following vaccination, which artificially increase the durability of their response. To roughly estimate the fraction of breakthrough infections, we found that only 1% of subjects did not reach their peak HAI at 1 month post-vac, but instead had a ≥4x larger HAI titer at day 75 or day 90. This suggested that there were few cases of a strong infection signature that would skew the HAI dynamics.

Note that the 2022 UGA study had a markedly larger peak fold-change at 1 month post-vac in all groups compared to the average across all other studies (FC_Weak,2022UGA_=2.3x vs FC_Weak,Avg_=1.6x, FC_Transient,2022UGA_=10.7x vs FC_Transient,Avg_=6.4x, FC_Durable,2022UGA_=33.3x vs FC_Durable,Avg_=16.3x; **Fig S5A**), and we assumed the 2x decrease described above from days 28-90 would hold in any other year. To emphasize that this assumption was based on a single dataset, we plot the day 90 responses with a dashed line (**Fig 1B, 4A**), and we tested this proposed trend as a prediction challenge as described in a subsequent section.

### Durable responses remain within ∼2x of their peak out to 1 year post-vaccination, whereas transient responses return to baseline

To characterize the long-term dynamics of the vaccine response, we analyzed fold-change from the 9 datasets where the 1 year post-vac response could be measured or inferred from the subsequent season (**Table 1**, white background). Whereas weak responders returned to their baseline fold-change of 1.0x (95% CI: 1.0-1.1x), transient responders decayed ∼2x from the peak to day 90 followed by another ∼2x decrease from day 90 to day 365, reaching a modestly-elevated fold-change of 1.5x (95% CI: 1.4-1.6x) (**Fig 5A**). Half of transient responders (50%) retained a 2x fold-change at 1 year post-vac, whereas 37% – similarly to weak responders – decayed to their 1x baseline (**Fig 5B,C**).

**Figure 5.**
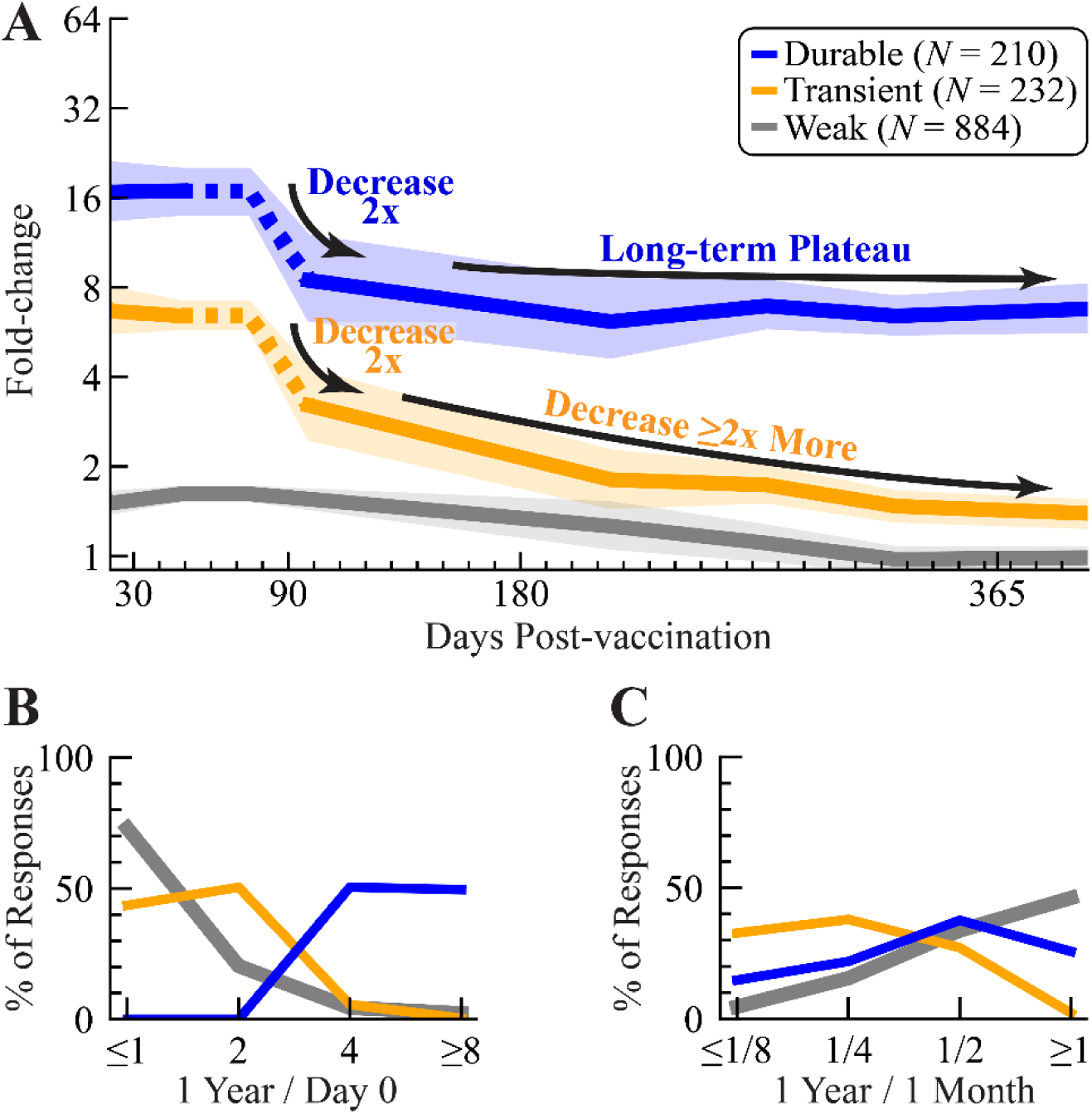
Timing and magnitude of the HAI titer response out to 1 year post-vaccination across durable, transient, and weak responders. (A) Fold-change for any participants measured between days 30-400 post-vaccination. Lines show the geometric mean fold-change and 95% confidence intervals. Dashed portions of these lines were derived from single datasets, in particular the 2010 Kennedy study^15^ for day 75 and the 2022 UGA study for day 90. (B) Comparing HAI titers at 1 year to day 0 or (C) at 1 year to 1 month post-vac.

Durable responders, by contrast, decayed ∼2-fold from the peak by day 90 and then maintained a fold-change of 6.8x out to 1 year post-vac (95% CI: 6.2-7.5x). Notably, 50% of durable responders exceeded the required cutoff (fold-change ≥4x at 1 year) and retained ≥8x fold-change (**Fig 5B**). Durable responders also sustained the greatest absolute HAI at 1 year, with a geomean of 131, compared to transient (geomean HAI=26) and weak responders (geomean HAI=48, weak responders had 2x higher day 0 HAI than transient and durable) (**Fig S1A**).

### Short- and long-term dynamics can be predicted within ∼2x with knowledge of an individual’s peak fold-change

Given the heterogeneity of responses ‒ for example, half of strong responders decay to baseline while the other half exhibit durable responses ‒ we tested how well a single measurement can predict the full response dynamics. That single measurement inferred whether an individual was weak (≤2x), transient (4-8x), or durable (≥16x when day 21 was measured; upper ranges 2x smaller for transient and durable when day 7 was measured, **Methods**). Using that classification, the remaining dynamics were extrapolated using simplified representations for each phenotype (**Fig 6A**), with root-mean-squared error (RMSE) quantifying prediction accuracy. As a point of comparison, we considered an “average model” replicating the traditional analysis that combines all subjects across all cohorts irrespective of their weak, transient, or durable phenotype (**Methods**).

**Figure 6.**
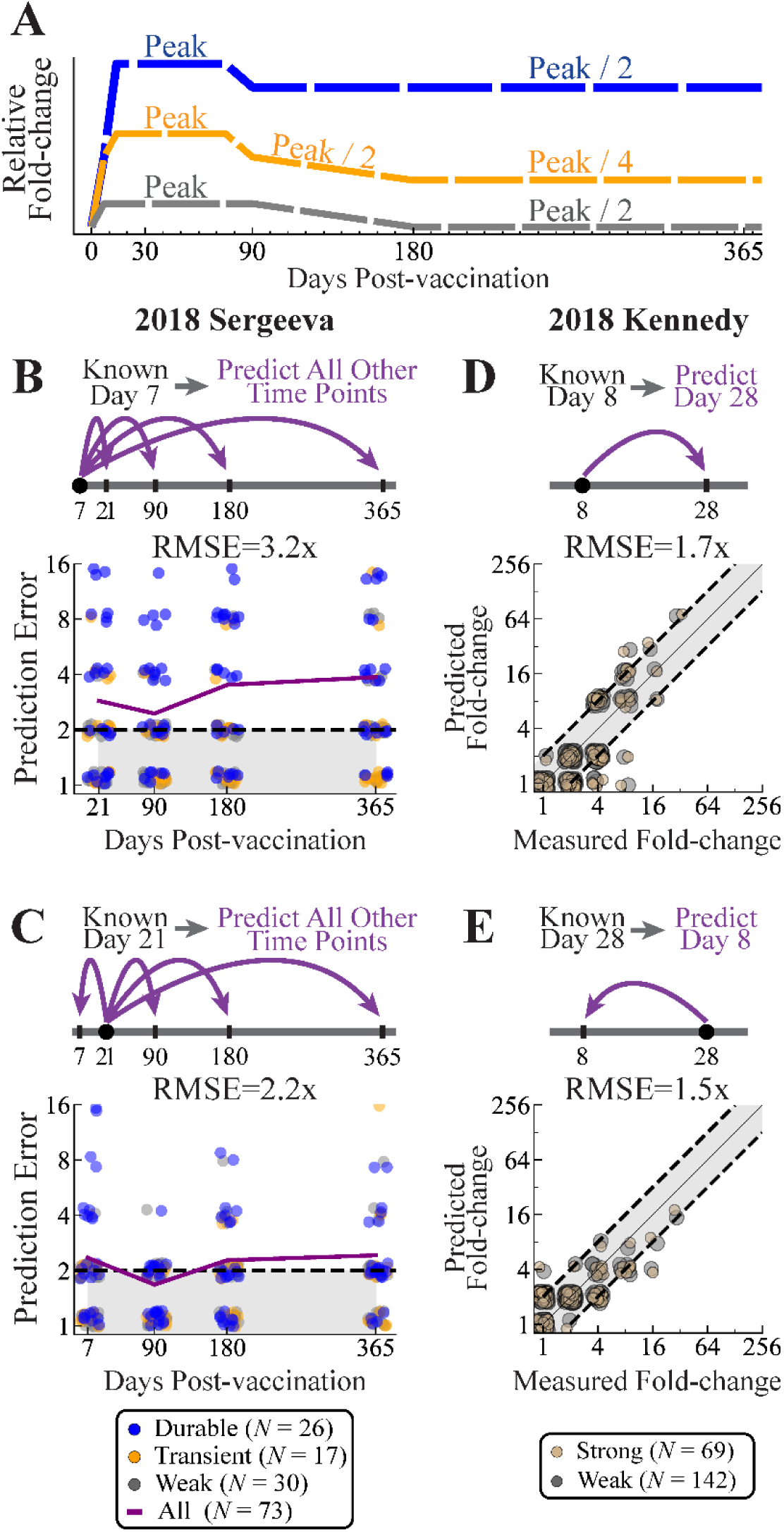
Using a single time point to predict both short- and long-term dynamics. (A) A simplified representation of weak, transient, and durable dynamics can be fit to a single time point to predict the full response. (B,C) Prediction RMSE across time points for 2018 Sergeeva using (B) only day 7 or (C) only day 21 fold-change.^16^ The purple line shows the RMSE at each time point. (D,E) Predicted versus measured fold-change for 2018 Kennedy using (D) only day 8 or (E) only day 28 fold-change. Points show individual predictions, with colors indicating their true category (weak, strong, transient, or durable), and not their inferred category used for fitting. Dashed lines surrounding gray shading represents the 2x error of the HAI assay.

We tested this approach using three studies with an increasing level of blindness. First, we searched for a new vaccine study, 2018 Sergeeva, that was not used in the prior analysis to define the HAI dynamics. This study measured vaccine responses (*N*=73) at days 0, 7, 21, 90, 180, and 365 against the H3N2 A/Singapore/INFIMH-160019/2016 vaccine strain.^16^ The computational team (A.L.) only used the earliest days 0 and 7 data, blinding themselves and predicting days 21, 90, 180, and 365 values (**Fig 6B**). Following that, the peak day 21 responses were used to predict the remaining time points (**Fig 6C**).

Second, we carried out a new 2018 Kennedy study (*N*=211), measuring HAI of banked sera against the H3N2 vaccine strain A/Singapore/INFIMH-160019/2016 at days 0, 8, and 28 (**Methods**). All subjects were ≥65 years old, and they were either given high-dose Fluzone or Fluad, with the same predictive framework applied to both vaccine formulations. The computational team was fully-blinded by the experimental team (R.K.), who only sent day 0 and day 8 data. Predictions were made and timestamped (see <u>GitHub</u>) before the day 28 measurements were provided for comparison (**Fig 6D**). Following this analysis, fold-change was then predicted in the opposite direction, using day 28 to predict day 8 (**Fig 6E**).

Third, we used samples available from the partially-complete 2023 UGA study (*N*=234) with HAI measured against the H3N2 A/Darwin/9/2021 vaccine strain as well as six historical variants (A/Hong Kong/4801/2014, A/Singapore/INFIMH-160019/2016, A/Kansas/14/2017, A/South Australia/34/2019, A/Hong Kong/2671/2019, and A/Tasmania/503/2020) that were chosen to overlap with the 2020-2022 UGA studies. At the time of writing, HAI was only measured for the day 0 and day 28 time points, with predictions timestamped (see <u>GitHub</u>) and awaiting the 1 year measurements (that will also be posted on GitHub when they are available).

In the first prediction challenge for 2018 Sergeeva, the RMSE across all predicted time points was 3.2x using day 7 HAI (**Fig 6B**), which was significantly smaller than predictions found by averaging all responses (RMSE_Avg Model_=4.4x, *p*<0.05, one-sided permutation test). When day 21 was known, RMSE across all predicted time points improved to 2.2x, mostly because the durable responses were predicted with ∼2x accuracy at later time points (**Fig 6C**), and these predictions remained significantly better than the average model (RMSE_Avg Model_=4.3x, *p*<0.05). Notably, the 2x drop from day 28→90 observed for transient and durable responders in 2022 UGA was also seen for most 2018 Sergeeva subjects (**Fig S4I**), corroborating that day 90 marks the end of the peak HAI response.

In the second challenge for 2018 Kennedy, predictions from day 8 to day 28 were highly accurate (RMSE=1.7x, **Fig 6D**), with slightly better accuracy in the reverse direction (RMSE=1.5x, **Fig 6E**). As before, splitting responses into strong/weak phenotypes performed slightly but significantly better than using an average response model in both the forward (RMSE_Avg Model_=2.0x, *p*<0.05) and reverse directions (RMSE_Avg Model_=3.4x, *p*<0.05). Moreover, the fraction of subjects with weak responses were similar between high-dose Fluzone (70%, *N*=110) and Fluad recipients (68%, *N*=101), as were their dynamics.

The first two prediction challenges demonstrate that the dynamics of the vaccine response are highly reproducible, that early time points can predict later responses with 2-3x accuracy based on HAI data alone, and that stratifying individuals into weak, transient, or durable responders improves prediction accuracy. Although the third prediction challenge cannot yet be verified, the actual measurements will be posted once they are available.

## Discussion

*“The dynamism of titers complicates our understanding of their protective effect, for they may change substantially over the course of a single study.” – Zhao et al.*^7^

While the heterogeneity in influenza H3N2 vaccine responses is well-recognized, many studies report the average response across a cohort and compare this average across seasons and subgroups. Moreover, responses have traditionally been observed at two time points, namely, pre-vaccination and 1 month post-vaccination. In contrast, by examining individual trajectories, we found greater heterogeneity *within each cohort* rather than across cohorts for both response magnitude and durability, suggesting that person-specific factors often outweigh season-specific effects.

Given the inherent 2x error of the HAI assay (*i.e.*, we expect 40% of HAIs to be accurate, 50% to be 2x off, and 10% to be 4x off), and that most fold-change lies between 1-8x, many responses may be incorrectly classified using thresholds such as fold-change≤2x or fold-change≥4x. We addressed this issue by requiring both the vaccine strain’s HAI and another variant’s HAI to achieve fold-change≥4x for strong responders, since the HAI of recent variants closely tracks that of the vaccine strain. At minimum, measuring the HAI of the current and prior vaccine strains not only provides the long-term time point for repeat vaccines, but also flags potential outliers.

Using vaccine strain and variants HAI titers, we categorized individuals as weak, transient, or durable responders based on their fold-change at 1 month and 1 year post-vac relative to baseline. We found that 67% of subjects exhibited weak responses, with 10/14 vaccine studies reporting ≥50% weak responders.^9,12–15^ This implies that most vaccine responses elicit ≤2x fold-change at all time points, which is within the noise of the HAI assay. In stark contrast, durable responders not only achieved ∼16x fold-change at 1 month post-vac, but maintained a 6.8x response out to 1 year post-vac.

A small post-vac fold-change is often observed when subjects have large pre-vac titers, yet this “antibody ceiling” effect only dominates for very large pre-vac HAI^15,17,18^. More precisely, while pre-vac HAI≥160 essentially guarantees a weak response, only 21% of individuals had such large titers. Individuals with smaller pre-vac HAI could be weak or strong, although there was always a tendency towards weak responses. For example, a subject with pre-vac HAI=40 had an approximately 20% chance to be durable, 20% chance to be transient, and 60% chance to be weak. Age had a smaller effect, while both sex and high vs standard-dose vaccination did not meaningfully affect the odds of a weak response.

Furthermore, we found no shared characteristics among the 4 studies with <50% weak responders. A unique feature of the 2010 Kennedy was that it only contained elderly individuals (ages 50-74, administering the vaccine Fluarix)^15^, although the new 2018 Kennedy study introduced in this work also examined elderly individuals (ages 65-91, administering high-dose Fluzone or Fluad) but found 62% of high-dose Fluzone and 73% of Fluad recipients had weak responses. Indeed, we expected the opposite results, given that high-dose Fluzone and Fluad are designed to elicit stronger responses in the elderly. 2016 UGA and 2016 Fox_Nam_ also had <50% weak responders, the former being the first in the series of UGA studies (where every subsequent year had 57-81% weak responders), while the latter represents a study in Ha Nam where adults were given the first influenza vaccine of their life. These two studies suggest that prior vaccination history may play a role in the fraction of weak responders. 2018 Sergeeva additionally had <50% weak responders, with no apparent special inclusion criteria, methodology, or host factors to directly explain this lower fraction. Note that in addition to neglecting vaccination history in our analysis, we also did not consider infection history and the potential impact of both influenza and other viral infections.^9,19^

In addition to the peak 1 month fold-change, the dynamics of the weak, transient, and durable responses also differed in several key ways. Whereas weak responses hovered around baseline, both transient and durable responses had a pronounced peak, with the former returning nearly to baseline by 1 year while the latter remained elevated. This suggests two immunological thresholds: one for starting a response (weak→transient) and another for mounting a lasting response (transient→durable).

These thresholds may reflect B cell activation thresholds, where weak responders fail to generate sufficient germinal center reactions, while durable responders recruit long-lived plasma cells to the bone marrow.^6^ Indeed, antibodies injected into humans have a half-life of 2-3 months^20^, similar to the decay seen in transient responders, suggesting that these transient responders generate a burst of antibodies that cease once the vaccine antigens are cleared. However, plasmablasts produce antibodies for >10 years^21^, and T cell help^22^ or platelet-associated factors^23^ may differentiate between the short-term and long-term production of antibodies. Our study is limited in that it only uses serum HAI as a readout, and future work will search for such mechanisms.

By splitting off the transient and durable responses from the weak responses, we explored the dynamics of the vaccine response with unprecedented resolution. We found that HAI began increasing 6-7 days post-vac, faster than the 10-15 days required for an antibody response to the SARS-CoV-2 vaccine in naive individuals^24,25^, likely because the children and adults in these studies were not influenza naive. Furthermore, the distinction between transient and durable responses already starts to emerge by days 6-7, where durable responses are ∼2x greater on average. A small vaccine study showed that in addition to HAI, binding of IgG, IgA, and IgM also increased 7-8 days post-vaccination [Table S2 of Henn *et al*.^14^].

Surprisingly, the peak influenza response was achieved by day 10 and lasted until day 75, providing a far longer plateau than expected. These results suggest that strong responders benefit from ∼2 months of a peak HAI response, and it calls to question whether studies measuring the peak response have chosen the 1 month time point based on historical precedent. Measuring the response earlier could not only simplify experimental logistics but could also better sample the immune response. For example, since T cell responses approach their peak ∼14 days post-vac^26^, that time point could more comprehensively assess both humoral and cellular immunity.

These dynamics are mostly in line with prior cohort-level analyses. A 1992 study of 68 subjects showed little-to-no fold-change at day 4, had a small response by days 6-7, and reached peak fold-change by days 14-15.^27^ A 2009 study found a near perfect match between antibody responses at day 7 and day 70 for all vaccine components (Fig S4 of Tsang *et al*.).^28^ A 2005-2006 study of 940 college students surprisingly found that their average response resembled the durable phenotype, but their dynamics matched those reported here, with fold-change of 8x at day 30 and 4x at day 365.^29^ A 2011 study of 160 subjects measured at 30 days and either 90 or 180 or 270 or 365 days post-vac found that H3N2 titers at day 30 were 2-4x larger than the later time points, although a 2003 study found a surprisingly flat response at all time points (Fig S1 of Plant *et al*.).^30^ Each of these studies lists average cohort responses based solely on the vaccine strain, and hence we cannot separate the weak, transient, and durable responders.

By using these three phenotypes, HAI measured at a single time point (as early as day 7, the earliest time point tested) could predict the titers out to 1 year post-vac significantly more accurately than an average model based on all responses, and these phenotypes could feed into computational approaches to predict person-level responses.^10,31^ These results are also in line with systems immunology approaches that used early vaccine responses (days 1-7 post-vac) to predict the peak response, and they can complement such efforts to show how much more information is gained by adding transcriptomics.^11,32^

One proposed application of such systems immunology approaches is to use early vaccine responses to identify individuals whose immunity would wane later in the season and give them a booster dose. Other work has suggested that a 2-dose vaccine, with a booster dose administered 3 months after a priming dose, could be especially effective in seasons where peak influenza cases occur late.^33^ However, once strong responders achieve their peak HAI at day 10, they stay within ∼2x of that peak response out to 4 months post-vac (the duration of the influenza season), and hence there may be minimal benefit in a booster dose if it elevates the titers back to their peak level.

Indeed, influenza boosters administered 1 month post-vac across multiple subgroups have elicited little-to-no increase in HAI^34–38^, and we expect at most a 2x response if the booster is given 3 months post-vac. Instead, there would be far more benefit in identifying a mechanism that could identify the majority of weak responders each season or turn them into strong responders. For example, different vaccine formulations^39,40^ (*e.g.*, high-dose Fluzone, Flublok, Fluad), adjuvants^41^, or next-generation vaccines^42^ have been proposed to serve that function, although our analysis of high-dose Fluzone and Fluad did not find that they elicited substantially larger responses.

Eliciting durable immunity against non-systemic respiratory viruses such as influenza, RSV, or SARS-CoV-2 remains an open challenge.^43^ Creating a durable vaccine response that lasts for multiple seasons (ideally for an entire lifetime) is one of the stated goals of universal influenza vaccines, together with increasing its breadth and effectiveness.^44^ We speculate that these goals of increased durability, breadth, and effectiveness may be somewhat complementary; next-generation vaccines with greater breadth may still need to develop mechanisms that increase durability, and the lessons learned from seasonal vaccines will likely carry over. For example, vaccines aimed at eliciting hemagglutinin stem-targeting antibodies may need to overcome the antibody ceiling effect^15,17^, and any durable vaccine will need to recruit long-lived plasma cells to the bone marrow.^6^ That said, classic work showed that whole virus can elicit durable responses out to 2 years post-vaccination, suggesting that the durability can be achieved with the right trigger to the immune system.^45^

At present, few vaccine studies measure long-term durability, and only 3/14 studies in this work measured such time points. Studies that repeatedly vaccinate the same group of individuals become far more informative, since a person’s long-term response is automatically determined (using next season’s pre-vac titers) when the vaccine does not change, and can be measured with minimal effort when the vaccine strain does change. Moreover, tracking the same individual across seasons will quantify whether individuals tend to maintain or change phenotypes, or whether more frequent vaccinations skew durable responses into transient or weak responses.^10,29,46,47^ Conversely, recent prior infections may turn a weak response to transient or durable.^9^

Taken together, these results suggest that current seasonal influenza vaccines consistently elicit weak responses across a large fraction of the population. This holds across age groups, seasons, and even the high-dose and adjuvanted vaccines studies we investigated. Roughly 33% of participants achieved a peak fold-change≥4x, comparable to the level of vaccine effectiveness across seasons, yet even these responses substantially reduce hospitalizations and deaths.^48,49^ Rather than providing cohort-level responses, this work calls for reports of individual vaccine responses, and for studies that specifically focus on both the weakest responders that are currently underserved by seasonal vaccines, and the strongest responders that demonstrate the immense utility of what vaccines can offer.

## Data Availability

To expedite the review process, the raw data and predictions will be attached as a CSV file. Upon publication, all data will be made available on GitHub.

## Acknowledgements

The authors acknowledge the participants in the UGA and Florida studies and the team members for sample collections blood and saliva processing and technical assistance including, Alexandra Abu-Shmais, Julia Aguirre, James D. Allen, Brittany Baker, Jordan Byrne, Jasmine Burris, Michael A. Carlock, Noah Cecil, Patrick Fagan, Brianna Gamez, Omar Hamwy, Hannah B. Hanley, John Ingram, Lauren Howland, Erin Jarrett, Hana Ji, David Lee, Mitchell Lee, Katie Mailloux, Zachary McGuire, Hua Shi, Frankie Stewart, Naoko Uno, Terris Wimbs, and Emma Whitesell. We would also like to thank all participants enrolled in the study, as well as Dr. Brad Phillips, Kimberly Schmitz, and the entire staff at the University of Georgia Clinical and Translational Research Unit (CTRU) for assistance in collecting samples in the influenza vaccine program.

This research was supported by the National Center for Advancing Translational Sciences of the NIH (CTRU, Award #UL1TR002378), Collaborative Influenza Vaccine Innovations Centers (CIVICs) of NIAID (TMR, Contract #75N93019C00052), University of Georgia (TMR), Cleveland Clinic (TMR), and by the Georgia Research Alliance (TMR, Georgia Eminent Scholar GRA-001).

## Methods

### Datasets analyzed

All vaccine studies (except the 2018 Kennedy and 2022 UGA studies introduced in this work and discussed below) are described in the following manuscripts: Henn 2013, Hinojosa_V_ 2020, Fox 2022, Carlock 2024, Jacobson 2015, and Sergeeva 2023.^9,12–16^ In the 2010 Kennedy, 2010 Henn, 2014 Hinojosa_V_ [Fluzone only], 2016 Fox_HCW_, and 2016-2019 UGA [high dose only] studies, trivalent vaccines were administered while all other vaccines in **Table 1** were quadrivalent. Note that in the UGA studies, the standard-dose Fluzone from 2016-2021 and the high-dose Fluzone vaccine from 2020-2021 were quadrivalent. In 2014 Hinojosa_V_, it was reported that 32 children received Fluzone while 9 received FluMist, and that 3 Fluzone recipients and all 9 FluMist recipients reported an H3N2 infection during the 2014 season, yet explicit subject IDs were not given; all subjects were included in our analysis.

The explicit time points measured in each study subtly differed. 2016 Fox_Nam_, 2016 Fox_HCW_, 2018 Sergeeva, and 2016-2017 UGA measured the response 21 days post-vac, whereas all other vaccine studies measured the day 28 post-vac response. In 2020-2022 UGA, the precise date of measurement was reported (*i.e.*, participants were asked to return 28 days post-vaccination, but the exact dates ranged from 21-71 days, and a similar spread is expected in all other studies). Such subtleties in timing were ignored in this analysis, and when a study measured the response between 13-60 days post-vac, the nearest time point to day 28 was treated as “1 month post-vaccination.” Similarly, when a measurement was taken between 180-500 days post-vac, the nearest time point to day 365 was treated as “1 year post-vaccination.”

### 2018 Kennedy and 2022 UGA Vaccine Study Participants

The 2018 Kennedy study recruited 221 individuals ages 65-91 from southeastern MN, who were given either high-dose Fluzone or Fluad. More specifically, 114 participants were given the 2018-19 high-dose [60 μg/component] Fluzone Quadrivalent (Sanofi Pasteur) vaccine containing H1N1 A/Michigan/45/2015 X-275, H3N2 A/Singapore/INFIMH-160019/2016 IVR-186, B/Maryland/15/2016 (a B/Colorado/6/2017-like virus, B/Victoria lineage), and B/Phuket/3073/2013 (B/Yamagata lineage), while the remaining 107 participants were given the 2018-19 [15 μg/component] Fluad Trivalent (Seqirus) vaccine containing MF59 adjuvant and the aforementioned viruses except B/Phuket/3073/2013 (B/Yamagata lineage). Both the high-dose Fluzone and Fluad groups were jointly analyzed in this work, but predictions were only cast for the *N*=211 subjects with measurements at both days 8 and 28.

The 2022 UGA study measured 245 individuals from medical facilities near Athens GA, though in this work we selected 168 individuals who participated in the following season’s vaccine study to provide a long-term time point. Participants were given the 2022-23 vaccine comprising H1N1 A/Victoria/2570/2019, H3N2 A/Darwin/9/2021, B/Austria/1359417/2021 (B/Victoria lineage), and B/Phuket/3073/2013 (B/Yamagata lineage). As in prior UGA studies, participants less than 65 years old were given standard-dose [15 μg/component] Fluzone Quadrivalent (Sanofi Pasteur). Participants aged 65 or older were offered the high-dose [60 μg/component] Fluzone Quadrivalent (Sanofi Pasteur), and 3/5 of participants in this age group opted for the high-dose vaccine (see the supplemental dataset for information on vaccine dose).

### Ethics Statement

The 2018 Kennedy study was given ethical approval by the Mayo Clinic Institutional Review Board, and the 2022 UGA study was given ethical approval by the Western Institutional Review Board and the University of Georgia Review Board. Both studies received formal, written consent from each participant.

### 2018 Kennedy and 2022 UGA Variants Measured

Two new vaccine studies were conducted in this work. The 2018 Kennedy study only measured HAI titers against the vaccine strain H3N2 A/Singapore/INFIMH-160019/2016; while prior work analyzed the average response for this cohort at day 28,^50^ this work also measured the day 8 responses and analyzed all responses with person-level resolution. The H3N2 vaccine strain was purchased from Microbiologics (San Diego, CA). Virus stock was propagated in Certified Specific Pathogen Free embryonated chicken eggs.

The 2022 UGA study measured HAI titers against H3N2 A/Hong Kong/4801/2014, A/Singapore/INFIMH-160019/2016, A/Kansas/14/2017, A/South Australia/34/2019, A/Hong Kong/2671/2019, A/Tasmania/503/2020, and A/Darwin/9/2021. All viruses were propagated in embryonated chicken eggs in the lab of Dr. Ted Ross.

### Computing the geometric mean and 95% confidence intervals across time

When plotting fold-change vs days post-vaccination using multiple datasets (*e.g.*, **Fig 1B** and **Fig 3A**), data were binned as follows: days 0, 4, 7, 14, 21, 22-75, 76-120, 180-250, 251-300, 301-350, 351-500, where the midpoint of each bin was used for plotting. Nearest neighbor smoothing (current bin weight=1.0, next and previous bin weights=0.25) was applied to reduce abrupt changes between adjacent bins. When single datasets were shown (*e.g.*, **Fig 2C)**, each of their measured time points was shown without binning. 95% confidence intervals were computed using bootstrapping, where each group was resampled with replacement *N*=1000 times, and the 2.5th and 87.5th percentiles of the bootstrap distribution defined the lower and upper bounds, respectively. The same nearest neighbor smoothing was applied to the upper and lower limits of the 95% confidence intervals.

### Categorizing weak, transient, or durable responses using the vaccine strain and variants

All sera in **Table 1** (white background) were measured against the vaccine strain and other variants pre-vac, 1 month post-vac, and 1 year post-vac. Sera were categorized as transient or durable if the HAI fold-change of the vaccine strain and any additional variant reached ≥4x at 1 month post-vac (and otherwise categorized as weak). The additional variant is used to ensure the vaccine strain’s HAI fold-change is a true signal and not noise, since recent variants tend to have similar HAI dynamics to the vaccine strain. No constraints were placed on which prior variant must reach ≥4x fold-change, and every prior study in **Table 1** included the vaccine strain and at least six prior variants.

The additional variant served the same purpose at the 1 year time point. For durable responses, the vaccine strain and any additional variant must also reach ≥4x at 1 year post-vac, while transient responders did not meet this criteria at the 1 year time point. Any sera within the datasets listed in **Table 1** (white background) that did not have both a 1 month and 1 year time point were excluded from our analysis.

Since the 2010 Henn and 2010 Kennedy studies did not include a 1 year post-vac time point nor measured HAI against any variants, sera were categorized using HAI fold-change against the vaccine strain only at 1 month post-vac (weak if FC_1 month_≤4x, and strong if FC_1 month_≥4x).

### Using single datasets to infer dynamics in combined datasets

The 2010 Kennedy and 2022 UGA were the only studies where we inferred that the peak antibody response lasts out to day 75 and wanes by 2x at day 90. However, since the 2022 UGA study had a 2x larger peak HAI than the average response from other studies, we extrapolated the dynamics in **Fig 1B** as a horizontal dashed line to day 75 from the average response followed by a 2x decay to day 90. These dynamics at day 90 were corroborated when predicting the 2018 Sergeeva results.

### Intrinsic noise of the HAI assay

Harvey *et al*. showed that across the ∼700,000 ferret HAIs used for influenza surveillance, repeat measurements for the same serum-virus pair were consistent with Gaussian error (on a log_2_ scale) with standard deviation *σ*=1 (i.e. 2-fold error).^51^ More precisely, 40.0% of their repeat measurements did not change (1x error), 44.7% had 2x error, 12.6% had 4x error, 2.4% had 8x, and 0.2% had 16x error. In comparison, a log-Gaussian error distribution would predict that 38.7% of measurements would not change, 48.3% have 2x error, 11.7% have 4x error, 1.1% have 8x error, and 0.03% have 8x error.

In addition, Fonville *et al*. analyzed HAI measurements from nearly identical sera and found that the inherent error of the assay is log-normally-distributed with standard deviation ≈2-fold.^52^ This is shown by Figure S8B in Fonville *et al* (neglecting the stack of not-determined measurements outside the dynamic range of the assay), where 40% of repeats had the same HAI value, 50% had a 2-fold discrepancy, and 10% had a 4-fold discrepancy.

### Quantifying prediction error

Prediction error was quantified in unlogged units so that it can be readily compared to the measured values. RMSEs were computed by first taking the root-mean-squared error *σ* of the log_10_(HAI titers) and then presenting the un-logged value that is exponentiated by 10 (*i.e.*, *σ*=0.3 for log_10_ titers corresponds to an error of *σ*_Predict_=10^0.3^=2-fold, with “fold” or “x” indicating an un-logged number).

### Using a single time point to predict the full response dynamics

In addition to the course-grained representations of the weak, transient, and durable responses shown in **Fig 5A**, we also used the single time point to determine the phenotype of the response. If the single measurement occurred during the peak response (days 10-75), an individual was classified as weak if their fold-change was ≤2x, transient if it was 4-8x, and durable for fold-change≥16x. When using an earlier measurement from 6-8 days post-vaccination, we used fold-change≤2x for weak, 4x for transient, and ≥8x for durable responses.

The “average” model used as a comparator for the 2018 Sergeeva and 2018 Kennedy prediction challenges shows the geomean fold-change across all individuals (*not* split by weak, transient, or durable) from the 2014-2021 seasons (**Table 1**, white background). Responses used the same **Fig 1B** bins described above. Since no training datasets measured day 90, the predicted fold-change at this time point was linearly interpolated.

## ASupplemental Materials

**Figure S1.**
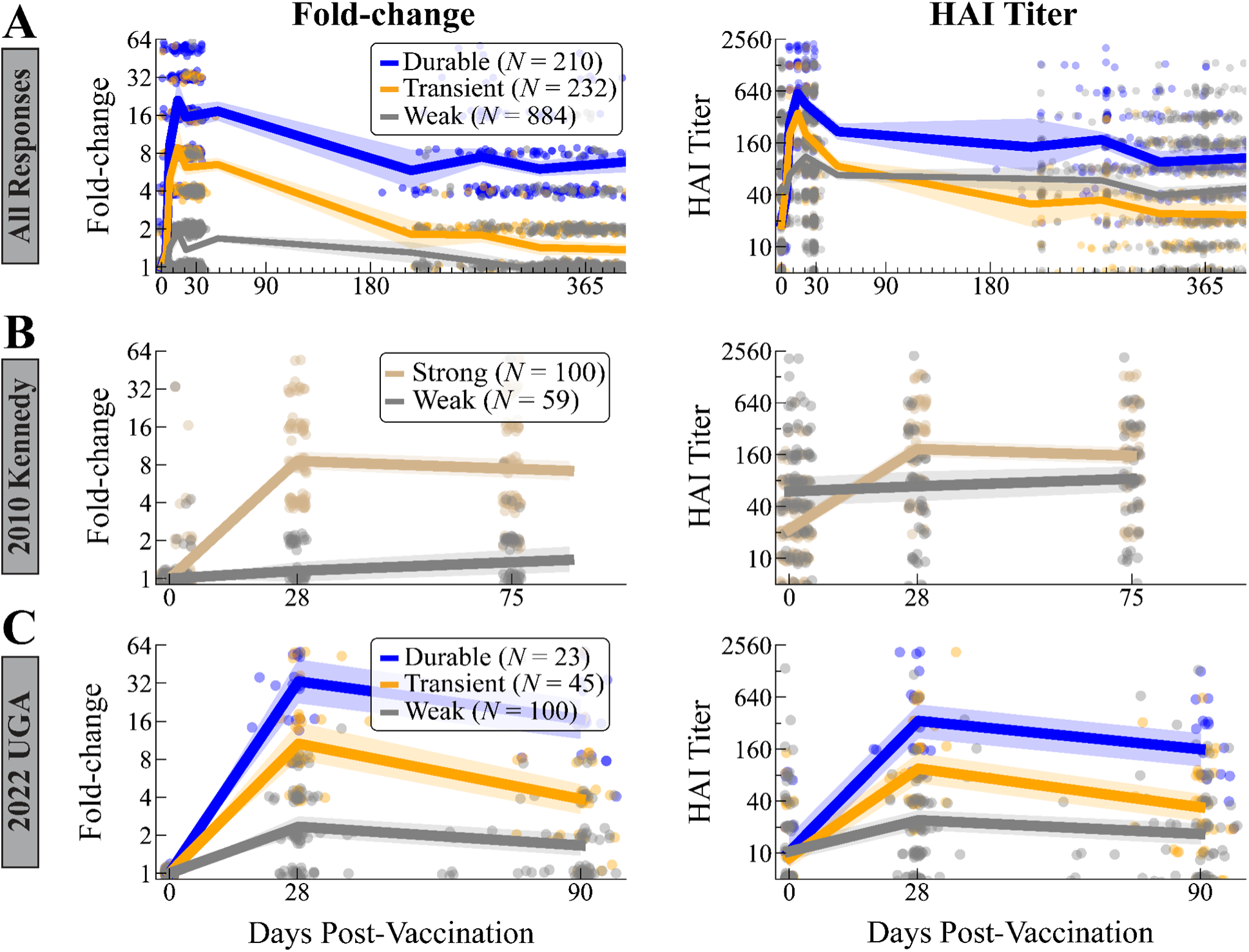
Comparing fold-change and absolute HAI titer. Fold-change [*left*] and absolute HAI titer [*right*] for durable, transient, and weak responders between (A) days 0-400 for all sera in **Table 1** (white background), (B) days 0-100 for 2010 Kennedy, and (C) days 0-100 for 2022 UGA. All plots are shown without the smoothing effects used in the main text. Legends apply to the left and right panels within each row.

**Figure S2.**
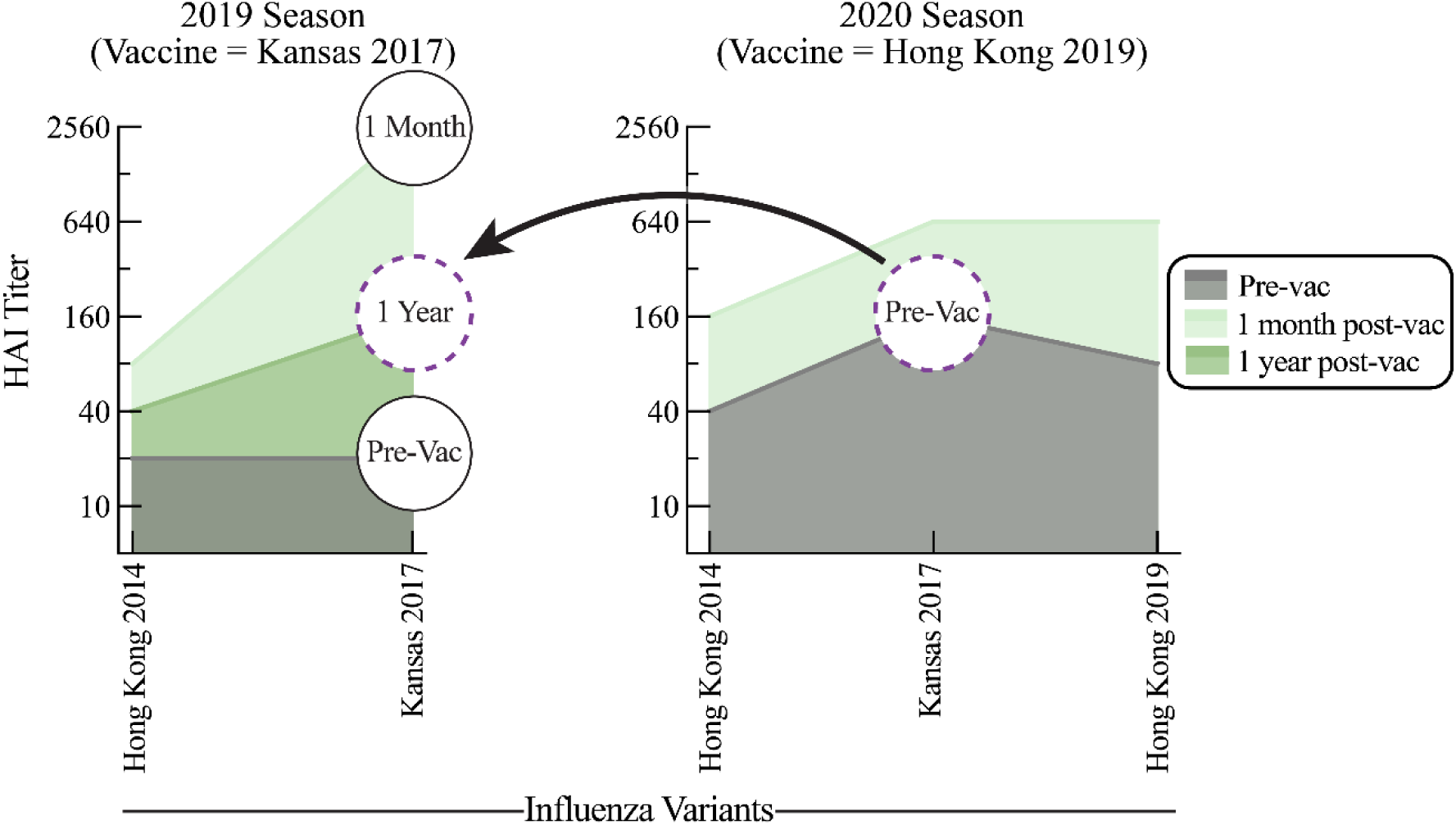
Pre-vac titers from the subsequent 2020 season determine the HAI titer 1 year post-vaccination from the 2019 season when prior vaccine strains are measured. Plots show hypothetical HAI titers from a single individual in a vaccine study carried out in both 2019 and 2020. Although the 2020 vaccine strain changed from H3N2 A/Kansas/14/2017 to H3N2 A/Hong Kong/45/2019, the pre-vaccination time point for H3N2 A/Kansas/14/2017 in 2020 provides the 1 year post-vaccination time point in the 2019 study. We assume the common scenario where each vaccine study measures current and prior vaccine strains, so that the 2019 study only measures H3N2 A/Hong Kong/4801/2014 and H3N2 A/Kansas/14/2017, whereas the 2020 study measures both prior vaccine strains and the current vaccine strain H3N2 A/Hong Kong/45/2019. This same method can be used across every season, and in seasons when the vaccine strain did not change (such as 2016-2017 and the 2017-2018 for H3N2), it can be used for studies that only measure the vaccine strain.

**Figure S3.**
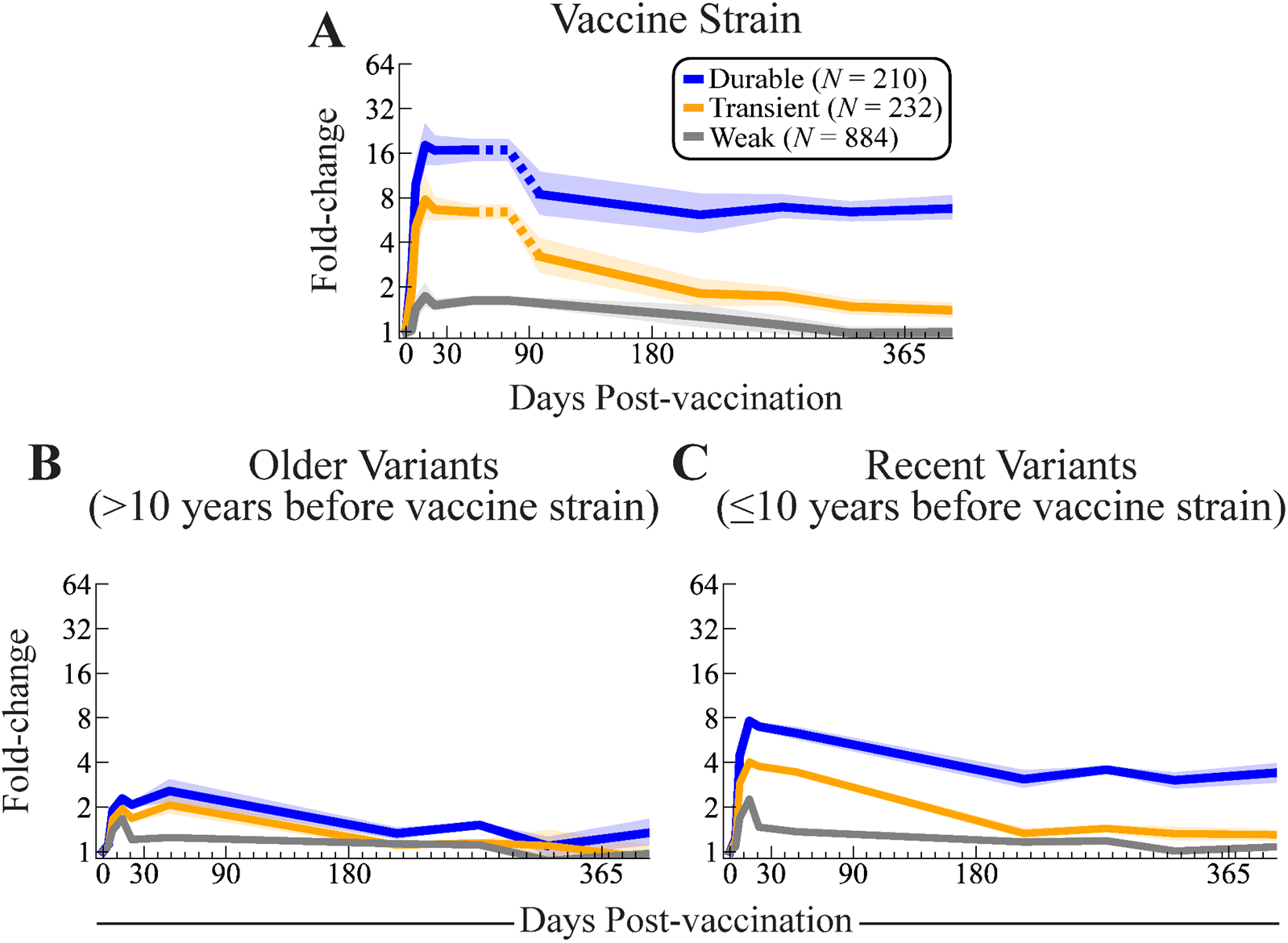
Long-term dynamics of the vaccine response against variants. Comparing the average vaccine response dynamics for durable, transient, and weak groups (A) against the vaccine, (B) variants circulating within 10 years of vaccination, and (C) variants circulating more than 10 years before vaccination.

**Figure S4.**
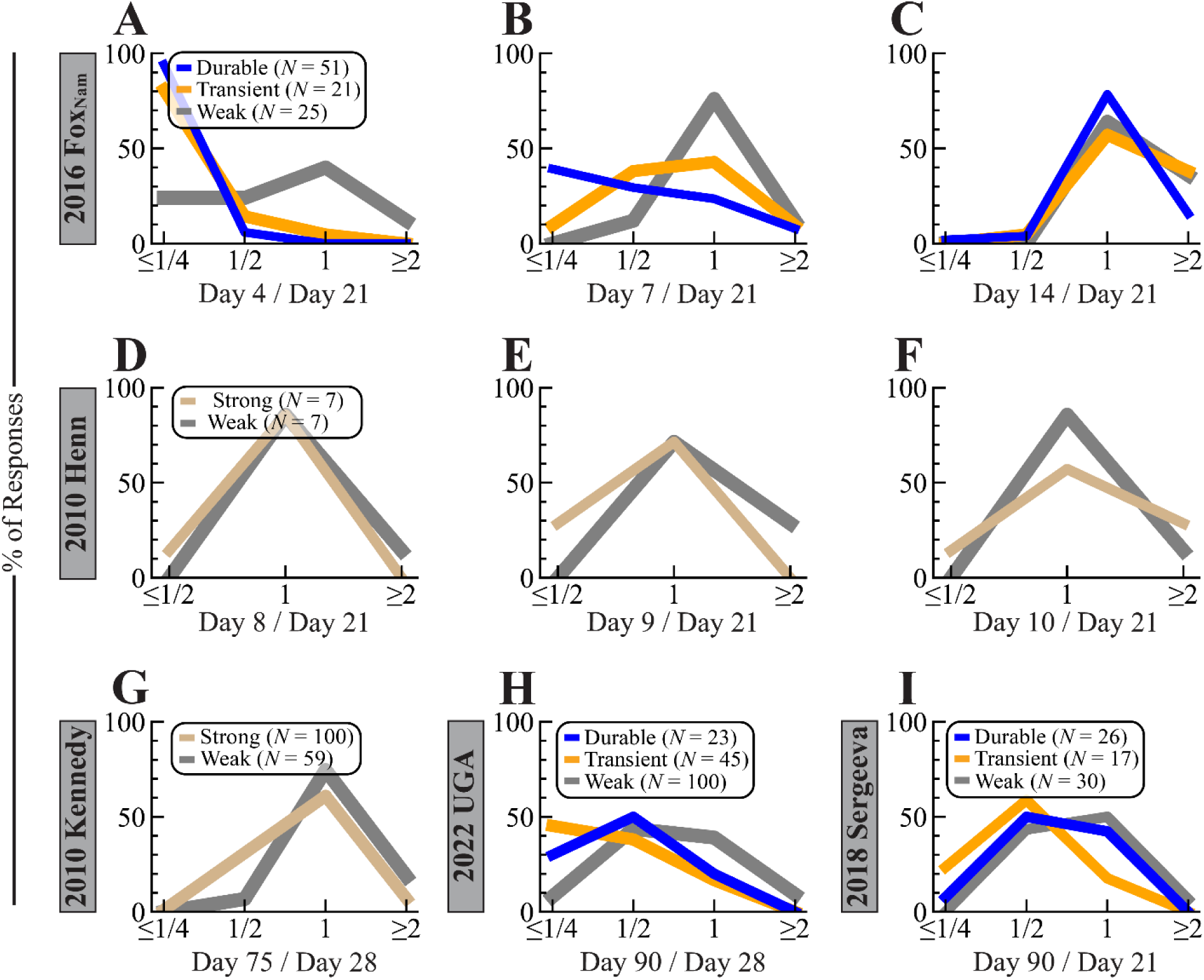
Comparing HAI titer at different time points in individual datasets used to determine peak timing. (A-C) Comparing HA titer against the vaccine strain (H3N2 A/Hong Kong/4801/2014) at early time points to day 21 within the 2016 Fox_Nam_ study.^9^ (D-F) Comparing HAI titer against the vaccine strain (H3N2 A/Perth/16/2009) at early time points to day 21 within the 2010 Henn study.^14^ (G) Comparing HAI titer against the vaccine strain (H3N2 A/Perth/16/2009) at days 28 and 75 within the 2010 Kennedy study.^15^ (H) Comparing HAI titer against the vaccine strain (H3N2 A/Darwin/9/2021) at days 28 and 90 within the 2022 UGA study.^13^ (I) Comparing HAI titer against the vaccine strain (H3N2 A/Singapore/INFIMH-160019/2016) at days 21 and 90 within the 2018 Sergeeva study.

**Figure S5.**
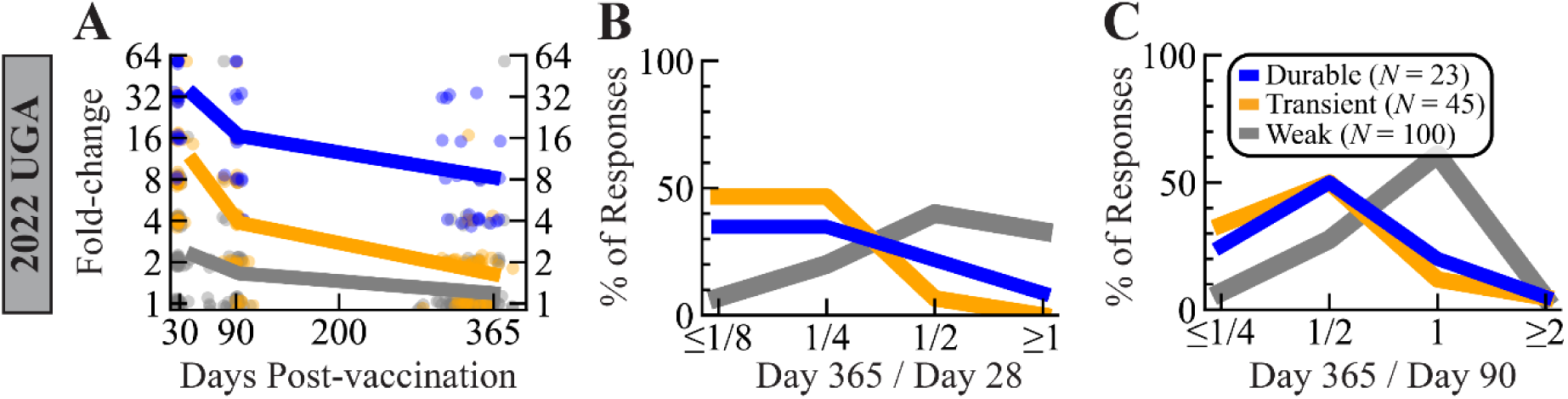
Long-term dynamics in the 2022 UGA study. (A) Fold-change against the vaccine strain (H3N2 A/Darwin/9/2021) at days 28, 90, and 365. HAI titer at (B) day 365 compared to day 28 or (C) day 365 compared to day 90. The number of individuals shown in the Panel C legend apply to all panels.

**Table S1.** Geometric mean HAI titers and fold-change across seasons at key time points. Statistics represent all subjects in each study. Fold-change is measured relative to pre-vac titers (and hence pre-vac fold-change=1 by definition).

| Influenza Season<br>(Studies from Table 1) | Pre-vac<br>GMT (FC) | 1 Month Post-vac<br>GMT (FC) | 1 Year Post-vac<br>GMT (FC) |
| --- | --- | --- | --- |
| 2014-15<br>(2014 Hinojosa<sub>v</sub>) | 183 (1.0x) | 348 (1.9x) | 304 (1.7x) |
| 2016-17<br>(2016 FoX<sub>Nam</sub>, 2016 FoX<sub>HCW</sub>,<br>2016 UGA) | 35 (1.0x) | 169 (4.7x) | 96 (2.7x) |
| 2017-18<br>(2017 UGA) | 100 (1.0x) | 199 (2.0x) | 74 (0.7x) |
| 2018-19<br>(2018 UGA) | 66 (1.0x) | 123 (1.9x) | 75 (1.1x) |
| 2019-20<br>(2019 UGA) | 11.7 (1.0x) | 53 (4.5x) | 20 (1.7x) |
| 2020-21<br>(2020 UGA) | 38 (1.0x) | 87 (2.3x) | 39 (1.0x) |
| 2021-22<br>(2021 UGA) | 32 (1.0x) | 73 (2.3x) | 52 (1.6x) |
| **Average** | **35 (1.0x)** | **102 (2.9x)** | **51 (1.5x)** |

**Table S2.** Studies without variants included in this work. This table lists studies that were included in this work despite not having variants. 2010 Henn and 2010 Kennedy provided unique time points that were absent from **Table 1** and were therefore included. 2018 Sergeeva also lacked variants but was used as an existing study to validate our predictive analysis.

| Year + Name of Study* | # Data Points | # Sera | # H3N2 Variants | Times Measured | H3N2 Vaccine Strain (Formulation) |
| --- | --- | --- | --- | --- | --- |
| 2010 Henn<sup>14</sup> | 168 | 14 | 1 | [0, 1, 2, 3, 4, 5, 6, 7, 8, 9, 10, 21] | A/Perth/16/2009 (FluLaval) |
| 2010 Kennedy<sup>15</sup> | 636 | 159 | 1 | [0, 3, 28, 75] | A/Perth/16/2009 (Fluarix) |
| 2018 Sergeeva<sup>16</sup> | 436 | 73 | 1 | [0, 7, 21, 90, 180, 365] | A/Singapore/INFIMH-160019/2016 (Ultrix, Grippol Plus, or Sovigripp) |

**Table S3.** Key statistics for the influenza H3N2 vaccine response. Separate statistics (and the number *N* of sera) are shown for the (A) pre-vaccination, (B) response initiation, (C) 1 month post-vaccination, (D) peak timing, and (E) 1 year post-vaccination responses. When datasets only measured responses out to 1 month post-vac, combined statistics are shown for transient and durable responders (collectively called strong responders). HAI fold-change (FC) is measured relative to pre-vaccination titers. As in Table 1, white backgrounds denote prior studies while gray backgrounds represent new studies introduced in this work.

| Magnitude of the Pre-vaccination Response |  |  |  |
| --- | --- | --- | --- |
| Year + Name of Study | Weak GMT | Transient GMT | Durable GMT |
| 2014 Hinojosa<sub>v</sub> | 210<br>[N=36] | 320<br>[N=1] | 48<br>[N=4] |
| 2016 FoX<sub>Nam</sub> | 47<br>[N=25] | 40<br>[N=21] | 28<br>[N=51] |
| 2016 FoX<sub>HCW</sub> | 49<br>[N=31] | 15<br>[N=10] | 24<br>[N=8] |
| 2016 UGA | 53<br>[N=47] | 28<br>[N=20] | 28<br>[N=35] |
| 2017 UGA | 125<br>[N=118] | 48<br>[N=20] | 38<br>[N=12] |
| 2018 UGA | 82<br>[N=131] | 25<br>[N=20] | 33<br>[N=11] |
| 2019 UGA | 14<br>[N=182] | 9<br>[N=80] | 10<br>[N=58] |
| 2020 UGA | 48<br>[N=196] | 19<br>[N=38] | 12<br>[N=16] |
| 2021 UGA | 40<br>[N=118] | 17<br>[N=22] | 13<br>[N=15] |
| **Average** | **48** | **18** | **19** |
| 2010 Kennedy | 60<br>[N=59] | 22<br>[N=100]<br>(strong) |  |
| 2010 Henn | 119<br>[N=7] | 27<br>[N=7]<br>(strong) |  |
| 2018 Sergeeva | 38<br>[N=30] | 18<br>[N=17] | 9<br>[N=26] |
| 2022 UGA | 10<br>[N=100] | 9<br>[N=45] | 10<br>[N=23] |
| 2018 Kennedy | 43<br>[N=142] | 26<br>[N=69]<br>(strong) |  |

(B)
| Days post-vaccination when HAI increases |  |
| --- | --- |
| Year + Name of Study | Days when strong responses reach $\geq 4x$ FC |
| 2016 FoxNam | Day 5-7<br>[N=72] |
| 2010 Henn | Day 6-8<br>[N=7] |
| Intersection | Day 6-7 |

(C)
| Magnitude of 1 Month (Peak) Response |  |  |  |
| --- | --- | --- | --- |
| Year + Name of Study | Weak GMT (FC) | Transient GMT (FC) | Durable GMT (FC) |
| 2014 Hinojosa<sub>v</sub> | 291 (1.4x)<br>[N=36] | 1280 (4.0x)<br>[N=1] | 1280 (26.9x)<br>[N=4] |
| 2016 FoX<sub>Nam</sub> | 66 (1.4x)<br>[N=25] | 280 (7.0x)<br>[N=21] | 559 (19.6x)<br>[N=51] |
| 2016 FoX<sub>HCW</sub> | 73 (1.5x)<br>[N=31] | 113 (7.5x)<br>[N=10] | 226 (9.5x)<br>[N=8] |
| 2016 UGA | 71 (1.3x)<br>[N=47] | 149 (5.3x)<br>[N=20] | 326 (11.7x)<br>[N=35] |
| 2017 UGA | 167 (1.3x)<br>[N=118] | 269 (5.7x)<br>[N=20] | 678 (18.0x)<br>[N=12] |
| 2018 UGA | 112 (1.4x)<br>[N=131] | 126 (5.1x)<br>[N=20] | 363 (11.0x)<br>[N=11] |
| 2019 UGA | 31 (2.2x)<br>[N=182] | 69 (8.0x)<br>[N=80] | 185 (19.1x)<br>[N=58] |
| 2020 UGA | 79 (1.6x)<br>[N=196] | 105 (5.6x)<br>[N=38] | 167 (14.1x)<br>[N=16] |
| 2021 UGA | 62 (1.5x)<br>[N=118] | 83 (5.0x)<br>[N=22] | 232 (17.6x)<br>[N=15] |
| **Average** | **77 (1.6x)** | **112 (6.4x)** | **313 (16.3x)** |
| 2010 Kennedy | 69 (1.2x)<br>[N=59] | 186 (8.6x)<br>[N=100]<br>(strong) |  |
| 2010 Henn | 145 (1.2x)<br>[N=7] | 390 (14.5x)<br>[N=7]<br>(strong) |  |
| 2018 Sergeeva | 52 (1.4x)<br>[N=30] | 136 (7.7x)<br>[N=17] | 220 (23.9x)<br>[N=26] |
| 2022 UGA | 24 (2.3x)<br>[N=100] | 95 (10.7x)<br>[N=45] | 340 (33.0x)<br>[N=23] |
| 2018 Kennedy | 66 (1.5x)<br>[N=142] | 134 (5.2x)<br>[N=69]<br>(strong) |  |

(D)
| Days post-vaccination when the peak response occurs |  |  |
| --- | --- | --- |
| Year + Name of Study | Days for Peak Start | Days for Peak End |
| 2016 Fox<sub>Nam</sub> | Days 7-14<br>[N=72] | - |
| 2010 Henn | Days 10-21<br>[N=7] | - |
| 2010 Kennedy | - | Days >75<br>[N=100] |
| 2022 UGA | - | Days <90<br>[N=68] |
| **Intersection** | **Days 10-14** | **Days 76-90** |

(E)
| Magnitude of 1 Year Response |  |  |  |
| --- | --- | --- | --- |
| Year + Name of Study | Weak GMT (FC) | Transient GMT (FC) | Durable GMT (FC) |
| 2014 Hinojosa<sub>v</sub> | 274 (1.3x)<br>[N=36] | 320 (1.0x)<br>[N=1] | 761 (16.0x)<br>[N=4] |
| 2016 FoX<sub>Nam</sub> | 59 (1.2x)<br>[N=25] | 75 (1.9x)<br>[N=21] | 210 (7.4x)<br>[N=51] |
| 2016 FoX<sub>HCW</sub> | 60 (1.2x)<br>[N=31] | 28 (1.9x)<br>[N=10] | 123 (5.2x)<br>[N=8] |
| 2016 UGA | 86 (1.6x)<br>[N=47] | 53 (1.9x)<br>[N=20] | 173 (6.2x)<br>[N=35] |
| 2017 UGA | 68 (0.5x)<br>[N=118] | 57 (1.2x)<br>[N=20] | 240 (6.3x)<br>[N=12] |
| 2018 UGA | 78 (1.0x)<br>[N=131] | 29 (1.2x)<br>[N=20] | 265 (8.0x)<br>[N=11] |
| 2019 UGA | 18 (1.2x)<br>[N=182] | 13 (1.5x)<br>[N=80] | 63 (6.5x)<br>[N=58] |
| 2020 UGA | 41 (0.9x)<br>[N=196] | 24 (1.3x)<br>[N=38] | 70 (5.9x)<br>[N=16] |
| 2021 UGA | 54 (1.3x)<br>[N=118] | 26 (1.6x)<br>[N=22] | 106 (8.0x)<br>[N=15] |
| **Average** | **48 (1.0x)** | **26 (1.5x)** | **131 (6.8x)** |
| 2018 Sergeeva | 29 (0.8x)<br>[N=30] | 25 (1.4x)<br>[N=17] | 96 (10.4x)<br>[N=26] |
| 2022 UGA | 12 (1.2x)<br>[N=100] | 14 (1.6x)<br>[N=45] | 85 (8.2x)<br>[N=23] |

